# General-purpose time-series foundation models enable sample-efficient transfer learning in retinal electrophysiology

**DOI:** 10.64898/2026.08.17.26360646

**Authors:** Hunter L. Porter, Cory B. Giles, Srividya Kottapalli, Jonathan D. Wren

**Affiliations:** Genes and Human Disease Research program, Oklahoma Medical Research Foundation, Oklahoma City, OK, USA; Department of Biochemistry & Physiology and OU Health Stephenson Cancer Center, University of Oklahoma Health Sciences Center, Oklahoma City, OK, USA; Neuroscience Program, University of Oklahoma Health Sciences Center, Oklahoma City, OK, USA

**Keywords:** electrophysiology, electroretinography, transfer learning, pattern ERG, full-field ERG, time-series, retinal biomarkers

## Abstract

Electroretinography (ERG) measures the functional response of distinct retinal cells to light, but was largely displaced by structural imaging in the 2000s. Standardization efforts by the International Society for Clinical Electrophysiology of Vision (ISCEV) began in the late 1980s, and collapsed the rich time-series traces into reproducible components and implicit times. Recent improvements in hardware (RETeval) and software (artificial intelligence) may increase the utility of ERG data. However, no ERG-specific foundation models exist, and there are not enough public datasets to train one.

We asked whether time-series foundation models (FMs) trained without ERG-specific pre-training could be adapted through transfer learning. Using two public datasets, PERG-IOBA (pattern ERG with ocular diagnoses), and LEOPs (full-field ERG focusing on Autism Spectrum Disorder, ASD), we interrogated how FMs could improve over smaller within-domain models. We measured the binary (healthy/typically developing vs any annotation) and multiclass (specific family/diagnosis) classification performance of both frozen and fine-tuned FMs, alongside custom autoencoder and multiscale models, using patient-aware splits for cross validation. We benchmark the same architectures against PTB-XL, a large 12-lead ECG corpus, as both a control for each approach and to explore scaling behavior. We show that 1) pre-trained FMs can reconstruct masked traces from all three datasets, 2) frozen and fine-tuned embeddings, especially combined with multimodal metadata through masked autoencoders, performed best on classification tasks. Performance on PTB-XL was maintained down to 300 records, comparable in size to the ERG datasets. We could not reproduce published classification performance on the ASD task.

Taken together, these results support general purpose foundation models as a practical approach to ERG analysis.

## I. Introduction

### Ocular Electrophysiology

Ocular electrophysiology is a powerful approach for measuring function in the central nervous system (CNS) ^1^. By using specific electrode placement and stimulus pairs, we can disentangle populations of multiple neuronal subtypes, including photoreceptors, bipolar cells, retinal ganglion cells (RGCs), and can measure conduction into the occipital cortex ^2^. One approach, pattern electroretinography (PERG), presents alternating patterns (e.g., checkerboards or hexagonal grids) in which constant-luminance pattern reversal isolates the cells involved in central vision ^3^. This contrasts with the more classical full-field electroretinography (ffERG) which measures responses from the entire retina using a range of stimulus protocols and signal processing steps to isolate components of interest (e.g. band-pass filters for oscillatory potentials) ^2^. The International Society for Clinical Electrophysiology of Vision (ISCEV) introduced standards for these stimulus protocols and for the signal components extracted for interpretation. These standards followed a broad exploration period in ERG during the 1980s and were intended to make recordings more comparable across researchers.

ERG proved useful for detecting inherited retinal diseases, such as retinitis pigmentosa (RP) and cone dystrophies ^4^, as well as retinal involvement in other conditions, including cancer-associated retinopathy and multiple sclerosis-associated optic neuritis ^5^. The assay still sees narrow clinical use but was largely supplanted in the 2000s by imaging methods such as optical coherence tomography (OCT) ^6^, which were faster to perform and easier to standardize. Recent developments make ERG worth reconsidering: easier-to-use instruments such as RETeval ^7^, Troland (Td) protocols enabling mydriasis-free measurements, and improved methods for data analysis. Deep learning models are particularly well positioned to exploit this opportunity, both in processing raw ERG traces and in integrating their functional perspective with widely used structural imaging modalities.

### The Promise of Artificial Intelligence

Over the last decade, artificial intelligence (AI) models have risen to prominence, enabled by advances in compute and the availability of large public datasets ^8^. Notably, the origins of modern AI lie in the perceptron, a system designed by Frank Rosenblatt in 1957 and modeled after biological systems, including the retina ^9^. AI and neuroscience have co-evolved, with parallels in ideas and language accompanying breakthroughs on both sides. For example, the neuronally inspired rectified linear unit (ReLU) helped address the vanishing gradient problem ^10,11^. The current trend in AI research is to build foundation models (FMs), pre-trained models that learn rich representations of a data modality ^12^. These models can then be fine-tuned into new domains and enable stronger inference and representation learning without the large datasets traditionally required. Transfer learning then adapts these FMs to new purposes, such as classification tasks for medical diagnoses ^13^.

Computer vision models such as the Vision Transformer (ViT) have transformed image processing ^14^, and form the backbone of models fine-tuned for ophthalmology, such as RETFound ^15^. This adaptation has yet to occur in retinal electrophysiology, likely because no public dataset approaches the scale required for domain-specific pre-training. Nonetheless, just as ViT can be adapted to OCT despite its broad training base on generic images, we hypothesized that time-series FMs can be adapted to ERG, enabling more generalizable inference than custom models trained on ERG alone. We therefore took two pre-trained time-series FMs, MOMENT and TimesFM, and adapted them to two recently published ERG datasets.

### Applying Time-Series Foundation Models to ERG

MOMENT (385M parameters) is a masked transformer model, trained to reconstruct time-series data from traces with patches removed across contexts including electrocardiography (ECG) and electroencephalography ^16^. TimesFM (200M parameters) is a decoder-only model, similar in architecture to GPT-style language FMs, trained largely on Google Trends and Wikipedia page views with no biomedical pre-training examples ^17^. We compared these against models trained exclusively within the ERG datasets, representing the conventional approach to applying ML to a new domain, and against standard ISCEV components and any state-of-the-art models available. For the custom models, we used bidirectional long short-term memory (BiLSTM) autoencoders and transformer-based autoencoders, two architectures with established records in time-series analysis ^18^.

### Approach and Hypotheses

We evaluated the usefulness of FMs in the absence of large domain pre-training corpora using two public ERG datasets: PERG-IOBA, comprising pattern ERG recordings across a range of ocular diagnoses ^19^, and LEOPs, comprising RETeval-derived full-field ERGs from a cohort annotated with autism spectrum disorder (ASD) status ^20^. To assess performance in less data-scarce regimes and to estimate scaling behavior in medical signals, we additionally used PTB-XL, a large 12-lead ECG corpus ^21^. Prior classification results exist for both ERG datasets, but are difficult to compare against directly. In PERG-IOBA, one report described perfect accuracy using a commercial chatbot presented with spreadsheets of data from 12 control and 12 macular dystrophy examples ^22^; while it is notable that an agentic system can perform the task at all, such models are not reproducible, and 24 samples do not establish the performance ceiling for this dataset. A more recent study explored hybrid machine learning approaches similar to our own, reporting a best mean binary AUC of 0.76 ^23^. For ASD classification in ffERG, variable frequency complex demodulation (VFCDM) with conventional machine learning algorithms has been reported to reach AUC of approximately 0.9 ^24^, and UMAP-derived features AUC of 0.98 ^25^. Neither the VFCDM nor FoundationalECGNet implementations are publicly available, and the UMAP-based analysis was performed on an earlier release of the ffERG data. We therefore benchmarked all models on out-of-fold predictions under nested cross-validation rather than a single held-out split, making performance directly comparable across datasets and architectures. Unlike ERG, the ECG domain contains many strong models owing to data availability, so we benchmarked against the FoundationalECGNet (FECGNet, 7.5M params) architecture ^26^, which we also adapted for ERG as an intermediate multi-scale model. FECGNet and the VFCDM models are not publicly available, so we reimplemented them from description alone.

We tested two primary claims. First, that time-series foundation models transfer usefully to ocular electrophysiology, and would exceed within-domain only deep-learning models. This entails three sub-claims: 1) that deep learning models can reconstruct ERG traces from masked/compressed inputs, showing they capture trace structure which we can use instead of derived clinical components. 2) that they match or exceed classification performance of models trained on extracted clinical components, implying they use information in the full traces that derived components discarded. 3) That FMs pre-trained on non-ERG time series match or exceed our best within-domain optimized models, implying their inductive biases and learned featurization generalize well into ERG analyses. Where a published state-of-the-art model existed for a dataset, we used its reported performance as the comparison standard rather than our own baselines.

Second, that these representations combine with other data to improve inference beyond what either provides alone. This entails that fine-tuning on within-domain data toward the classification target improves on frozen FM embeddings, and that FM embeddings fused with metadata covariates in a masked autoencoder outperform either signal or metadata alone, implying the model integrates information across modalities. We further asked whether learned representations support tasks beyond diagnostic classification, testing age regression alongside the classification tasks. Throughout, we evaluated each architecture on PTB-XL as a control, ensuring that failures on ERG reflected the modality, task, or dataset size rather than a misconfigured architecture.

Deep learning models could reconstruct masked ERG traces in all three datasets (Figure 1), confirming that the signal supports learned representation within domain. FM-based models matched or exceeded clinical-component baselines on all classification tasks except the multiclass LEOPs 446 (Figure 2), and TimesFM was competitive with our best within-domain models, despite TimesFM having seen no biomedical signals during pre-training. Fine-tuning via LoRA had mixed results compared to frozen embeddings; notably, MOMENT embeddings fine-tuned with LoRA showed the strongest performance on PTB-XL, exceeding our FECGNet reimplementation. Fusing FM embeddings with age, sex, and visual acuity covariates in a masked multimodal autoencoder (MMAE) produced the strongest PERG models, supporting our second primary claim. In the ASD task, MMAEs gave the best binary performance, while clinical components in a multi-layer perceptron gave the strongest multiclass accuracy. Age regression in all tasks was relatively weak. On PTB-XL, performance was maintained down to subsamples of approximately 300 records, comparable in size to PERG-IOBA, indicating that the sample efficiency we observed on ERG is a property of the transfer-learning approach rather than of these particular datasets.

Our results on LEOPs did not reproduce previously published performance. Under patient-grouped, nested cross-validation, our best models reached ROC AUC of 0.618 ± 0.036 (mean ± SE across 5 outer folds; MMAE finalist, 9-step 446 Td), substantially below the 0.9 and 0.98 reported for VFCDM- and UMAP-based approaches on this dataset. We found that this performance was driven substantially by demographic covariates: a logistic regression using only age and sex, evaluated under the identical patient-grouped nested-CV folds, reached AUC 0.589 ± 0.047 (9-step 446 Td) / 0.661 ± 0.033 (LA3), and we were unable to recover the published values under leakage-free evaluation. We report the evaluation conditions under which each published result does and does not reproduce, and discuss the mechanisms we tested and ruled out.

## II. Methods

**Table 1.** Dataset descriptives.

|  | PERG-IOBA | LEOPs (9-step@446Td) | PTB-XL |
| --- | --- | --- | --- |
| **N (records / patients)** | 336 / 304 (23 multi-visit) | 163 / 163 | 21799 / 18869 (2111 multi-visit) |
| **Age, years (mean ± SD, range)** | $37.1 \pm 18.3$ (4–86, n=336) | $13.4 \pm 4.6$ (6–27, n=163) | $59.5 \pm 16.8$ (2–89, n=21506) |
| **logMAR VA (mean ± SD, n)** | $0.33 \pm 0.48$ (n=317) | — | — |
| **Sex** | Female=176, Male=160 | Female=60, Male=103 | Male=11354, Female=10445 |
| **Binary distribution** | Normal=106 (31.5%)<br>Abnormal=230 (68.5%) | Control (TD)=86 (52.8%)<br>ASD (+ADHD)=77 (47.2%) | NORM=9514 (43.6%)<br>Abnormal=12285 (56.4%) |
| **Multiclass distribution** | Normal=106 (31.5%)<br>Retinal=180 (53.6%)<br>Neuro-ophth=34 (10.1%)<br>Other=16 (4.8%) | Control=86 (52.8%)<br>ASD=65 (39.9%)<br>ASD+ADHD=12 (7.4%) | CD=2325 (10.9%)<br>HYP=1308 (6.1%)<br>MI=5424 (25.4%)<br>NORM=9514 (44.5%)<br>STTC=2817 (13.2%) |

### Datasets and Study Populations

#### PERG-IOBA

The PERG-IOBA dataset comprises pattern electroretinography recordings from 304 participants annotated with 83 unique ocular diagnoses ^19^. Data were collected at the Institute of Applied Ophthalmobiology (IOBA) University of Valladolid, Spain, from 2003-2022. Diagnoses are additionally grouped into four families: normal, retinal involvement, neuro-ophthalmological disease, and other, the last consisting largely of toxicological and infectious conditions. Age and logMAR visual acuity are provided as participant-level metadata, along with a unilateral indicator specifying the affected eye in abnormal records. Participants contributed between one and four traces per visit, and a subset contributed multiple visits. Sampling rate is ∼1700 Hz for all time steps shown.

#### LEOPs

The LEOPs dataset comprises full-field electroretinography recordings acquired with the RETeval device ^20^. Recordings were collected at two sites (Australia and the United Kingdom). We could not locate the exact accrual period. Participants are annotated as typically developing (TD), ASD, or ASD with co-occurring ADHD (ASD/ADHD); the comorbid group is rare in this dataset. Recordings follow ISCEV protocols; we used the 446 Td flash intensity from the ISCEV 9-step protocol (see Data Preprocessing). Each participant contributed a single record. Iris contrast rating and age are available as covariates; other continuous variables are recorded only for ASD-labeled participants and were therefore excluded. Sampling rate is ∼1953 Hz for all time steps shown.

#### PTB-XL

PTB-XL comprises 21,799 12-lead ECG records organized into five diagnostic superclasses: conduction disturbance (CD), myocardial infarction (MI), hypertrophy (HYP), ST/T change (STTC), and normal (NORM), with more than 1000 unique patients annotated per class ^21^. Recordings were collected using Schiller AG devices between October 1989 and June 1996, curated by the Physikalisch-Technische Bundesantalt. Age is provided as a participant-level covariate. Raw data parsing was aided by WFDB ^27^. We used records after resampling to 100Hz.

No evaluation of fairness across demographic subgroups was performed. No prospective sample size calculation was performed, as this study uses existing datasets rather than newly collected data. PERG-IOBA and LEOPs are, to our knowledge, among the largest available public ERG datasets. We used patient-aware nested cross-validation to make efficient use of the data we had while avoiding overfitting. Our data scale analysis (Figure 6) gives some evidence the datasets are of reasonable size for the transfer learning approaches we used here.

### Data Preprocessing

#### PERG-IOBA

Participants had a range of 1-4 traces per visit, which were aggregated by averaging. Participants with multiple visits were forced into the same folds for training/validation/testing purposes. We explored alternative methodologies such as learning on all traces simultaneously, picking traces closest in Euclidean space to the mean for a participant-visit pair (medoid selection), and selecting the traces for a participant closest to the mean embedding of their class, and found averaging the most performant, except for in the fine-tuned FM approach where medoid was selected. For a small number of traces (5/336) where no trough was detected to assign the N35, P50 amplitude was instead measured using the pre-stimulus baseline per the ISCEV 2024 standard. Visual acuity was unavailable for 19 visits and was mean-imputed against the training fold to avoid leakage.

#### LEOPs

Records were averaged between eyes. We initially explored the ISCEV light-adapted 3 (LA3) protocol, and later switched to the ISCEV 9-step protocol’s 446 Td flash intensity due to its record as the most informative trace in other works.

#### PTB-XL

PTB-XL provides two choices for data download, an iterative wget protocol and a pre-zipped file. Initially, we explored the wget version and found it was missing samples after a few iterations, moving to the 21,799 sample .zip file version which all results now use. 34 records had an age of >200 years, a published sentinel value from the source data, and were excluded for age regression evaluation.

We tried multiple signal processing approaches, including those published in FoundationalECGNet, such as Daubechies 4 wavelet denoising and Morlet transforms, but discarded them for ERG experiments since they degraded performance.

### Prediction Task Definitions

Sample labels from each dataset were taken as-is, no rescoring or normalization was performed.

#### PERG

Binary class labels were inherited from either Normal (0), or any diagnosis at all (1). Multiclass was tested on the “Family” provided labels (Normal, Retinal, Neuro-ophthalmological, and Other). We also tried specific annotations with the 5 most abundant classes, but focused on families since we lose ∼36% of records in the 5 class approach. Age and logMAR visual acuity were used for regression testing.

#### LEOPs

Binary class labels were inherited from Typically Developing (TD, 0), or either ASD diagnosis. Multiclass labels split ASD and ASD+ADHD into separate categories, but the comorbid ADHD samples were very rare in the dataset. We decided to forego age prediction in the small ranged cohort that also included children, and could not use other continuous variables as they were only covered in samples labeled with ASD.

#### PTB-XL

Binary classes were annotated NORM (0), vs abnormal (1). The 5 superclasses PTB-XL annotated were used for the multiclass task (CD, conduction disturbance; MI, myocardial infarction; HYP, hypertrophy; STTC, ST/T change, and NORM). We trained age regression models in the same architectures.

### Model Families Explored

We compared several approaches for encoding and predicting from each dataset.

#### Naive

A control majority-class/mean predicting floor for each task.

#### Clinical features

extracted ISCEV/ECG clinical components. PERG consisted of N35, P50, and N95 amplitudes and implicit times extracted with a custom algorithm. ffERG traces consisted of pre-extracted a-wave and b-wave amplitudes and implicit times, and summed oscillatory potential amplitudes. PTB-XL consisted of NeuroKit2-derived timing and ST/T amplitudes, multi-lead voltage/Sokolow-Lyon ^28^.

#### Frozen FM with Linear and Multilayer Perceptron Probes

MOMENT-1-large and TimesFM-1.0-200m were used as-provided for frozen embeddings, and used to train a simple logistic/linear regression model in sklearn^29^. MOMENT embeddings were 1024 dims, while TimesFM embeddings are 1280 dims concatenated across channels (E.g. 1280 X n_channels). These were collapsed by PCA before logistic regression when the dimensionality was larger than the fold size to prevent memorization of training data. Linear and MLP probes were fit in sklearn, using LogisticRegression for binary classification linear probes, MLPClassifier for MLP probes, Ridge regression for age/VA linear probes, and MLPRegressor for age/VA MLP probes.

#### Masked Multimodal AutoEncoders (MMAE)

Multimodal fusion encoders learned on each task, implemented in pytorch ^30^. For PERG, this included age, sex, visual acuity, and the target labels (binary/multiclass) trained separately, along with a positive control of the provided “unilateral” label that specifies the affected eye, but only for abnormal traces. LEOPs consisted of age, sex, iris contrast rating, and model embeddings. Finally, PTB-XL included only age and sex. During training, we use independent Bernoulli masking with mask probability at 0.3, and a higher mask rate on the target label (0.5). During evaluation, the target modality is forcibly masked and all other covariates are used, except in PERG-IOBA where we report the results with “unilateral” also forcibly masked.

#### Fine-tuned Low-Rank Adaptation (LoRA) FMs

LoRA hypernetworks were trained either to aid in the reconstruction task (PERG/LEOPs), or directly toward the classification task (PTB-XL). The reconstruction fine-tuned FMs were evaluated using the same sklearn heads as described above, while the PTB-XL fine-tunes included a pytorch linear layer after the embedding layer for classification training to be done end-to-end, similar to the FoundationalECGNet objective described below.

#### Multiscale

Replication of FECGNet architecture – consisting of convolutional block attention modules, graph attention networks, and a Time-series transformer head, integrated into a learned fusion block, before equally weighted binary and multiclass loss. For reconstruction tests, we added a term for L1 reconstruction loss, and we also included ADASYN-based upsampling for rare classes implemented in imbalanced-learn ^31^.

#### FoundationalECGNet (PTB-XL only)

Multiscale model at the original paper’s scope (7.5M parameters). FECGNet pre-trains on three large datasets, and its method for integrating channel dimensions across modalities is not specified in sufficient detail to reproduce without access to the code, which is not publicly available. We therefore reimplemented the architecture from its published description. The original evaluation fine-tunes to a small 15-lead corpus in which approximately 100 samples form the test set, so we instead evaluated on out-of-fold predictions under cross-validation, making the comparison directly parallel to our ERG experiments. We further deviated from the published methodology by having the model also learn the multiclass task, as the hand-coded features underperformed the majority class floor.

#### Bi-LSTM AutoEncoders (AE, PERG only)

We initially tried using custom BiLSTM autoencoders for pattern ERG data. They are discussed, but were not competitive with stronger approaches and not explored in 9-step 446 from LEOPs nor in PTB-XL.

#### Other approaches

We additionally explored sample efficient architectures/augmentation strategies (Supervised contrastive learning and conditional variational autoencoders for synthetic generation) and smart featurization controls (ExBlock, ROCKET) for PERG to compare alternative approaches to FMs and custom embeddings. These were less performant and not included in the full nested-cv evaluation sweep. ROCKET and ExBlock were used as implemented in sktime ^32,33^.

### Published Comparison Methods

VFCDM-based approaches are not publicly available ^24^ and were reimplemented from their published description. The UMAP-based approach ^25^ provides public code, which we ran as released. We note that the original analysis used a subset of samples from an earlier release of the ffERG data, so absolute performance is not directly comparable to results reported here on the full dataset. Neither method produced expected results, so we instead benchmarked against the extracted clinical components.

### Cross-Validation Methodology

Our main evaluation suite focused on a nested 5-fold CV splitting. We split on patient ID for each dataset with sklearn’s StratifiedGroupKFold to generate 5 outer folds. Then, we split 3 inner patient-grouped folds for hyperparameter tuning. LEOPs only contained 1 record per participant, so no patient-grouping was necessary. For PTB-XL, we explored both a traditional train/val/test split (used for scaling explorations), and a GroupKFold on patientID on all folds except the held out test set.

### Hyperparameter Optimization

Hyperparameter ranges were initially explored using a train/val/test split in PERG and LA3 traces, with params tuned using Optuna^34^ for the val set, and only post-optimization would they be tested on the held-out samples. We used these runs to set the ranges for the nested CV sweeps, excluding values that were consistently poor on all splits. We searched seeds before starting training for sets that contained at least 2 samples of each class to ensure the selected “best parameters” were on models that had to be expressive in the classification task. For PTB-XL, we chose ranges from the other dataset rather than running the full 100 trial optuna pre-sweep. Nested CV hyperparameter sweeps used 10 trials to select the best parameters, and trials were pruned if they did not improve after N epochs.

### Shapley Analysis

Shapley attribution was assessed in two forms: computed over samples of the traces themselves using WindowSHAP^35^ for their contribution to binary classification, and computed over modalities for their effect on classification in MMAEs. For the time-series Shapley analysis, the attribution is performed over a frozen MOMENT linear probe using StationaryWindowSHAP evalauted against the medoid normal and abnormal sample. The traces are split into windows of length 15 time steps, using 20 random samples as the background set, and evaluated over 100 sampled coalitions of windows to estimate the attribution of each.

**Figure 1:**
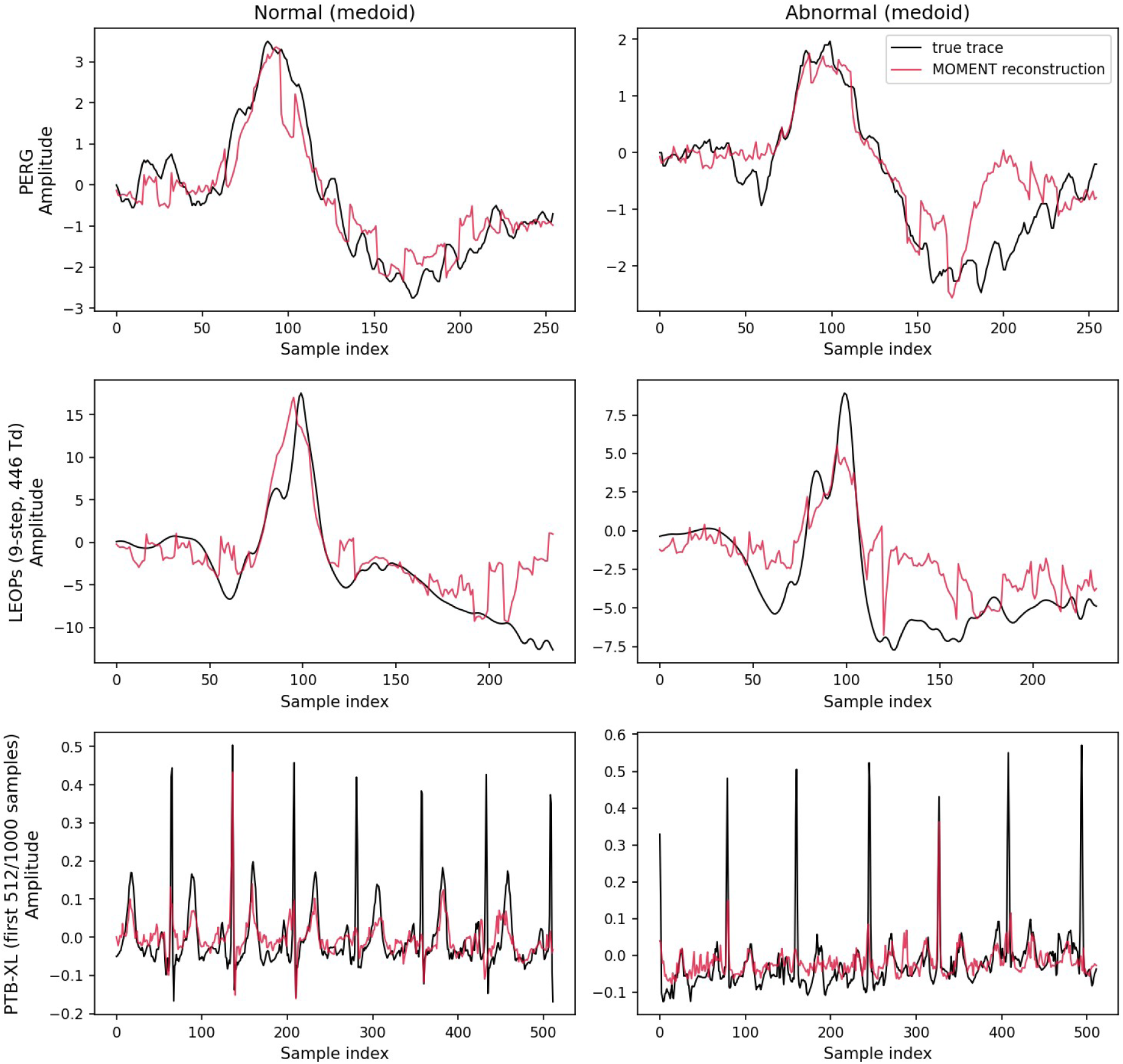
Signal reconstruction performance across modalities. Plots show masked reconstruction vs original trace for 1 example normal and abnormal trace from each dataset. PERG and LEOPs amplitudes in µV, PTB-XL in mV, sample index shows each time step.

For MMAE attribution, we used two separate approaches. For both ERG datasets, models were retrained using best hyperparameters while structurally removing modalities across all possible permutations, repeated across nested folds to obtain estimates of their value. For PTB-XL, we used post-training masking on all permutations of modalities to estimate their value against a fixed held-out test set.

### Statistical Analysis

All comparisons are reported as mean ± SEM. For each task and dataset, we computed an omnibus Friedman test across all finalists sharing that outer-fold split, followed by post-hoc comparisons for all pairs (Holm-corrected across the full family of pairwise tests) using both Wilcoxon signed-rank tests and paired t-tests. We used the paired t-test results as the primary post-hoc comparison, because analyses across n=5 folds for Wilcoxon returns a minimum p-value of 0.0625 regardless of effect size. Confidence intervals were derived using bootstrapping on out-of-fold results. PTB-XL’s scaling data was derived from resampling for fitting and tested against a fixed evaluation set instead, and the final point includes only one sampling and thus has no variance. All tests were using the SciPy stats ^36^, except Holm-adjusted t-tests which used scikit-posthocs ^37^.

### Evaluation Metrics

Binary performance was evaluated using primarily the area under the receiver operator characteristic curve (ROC AUC), alongside collapsed accuracy, F1 scores, and log loss. Multiclass models were scored on accuracy, macro-averaged F1 score, log loss, and one-vs-rest ROC AUC. Reconstruction was evaluated using masked L1 loss. Regression tasks were scored for both Pearson correlation coefficients and mean absolute error (MAE).

## III. Results

### Signal Reconstruction

Autoencoder and foundation-model architectures reconstructed ERG and ECG signals within domain across all three datasets. Reported losses are masked L1 reconstruction loss and are not directly comparable across datasets owing to differing raw-signal scales. Mean ± SE across 5 outer folds: PERG 1.20 ± 0.10, LEOPs (9-step, 446 Td) 4.21 ± 0.54, PTB-XL 0.114 ± 0.0002 (Figure 1).

On PERG (N=304, unsupervised, single split), Frozen MOMENT reconstructed PERG traces more accurately than any autoencoder trained from scratch under full visibility (MAE 0.336 ± 0.176) and, under genuine ∼50% patch masking (patches hidden and scored only on the hidden positions), still reconstructed accurately (masked MAE 0.546 ± 0.355); LoRA fine-tuning gave a small further improvement under masking (0.387 ± 0.221).

**Figure 2:**
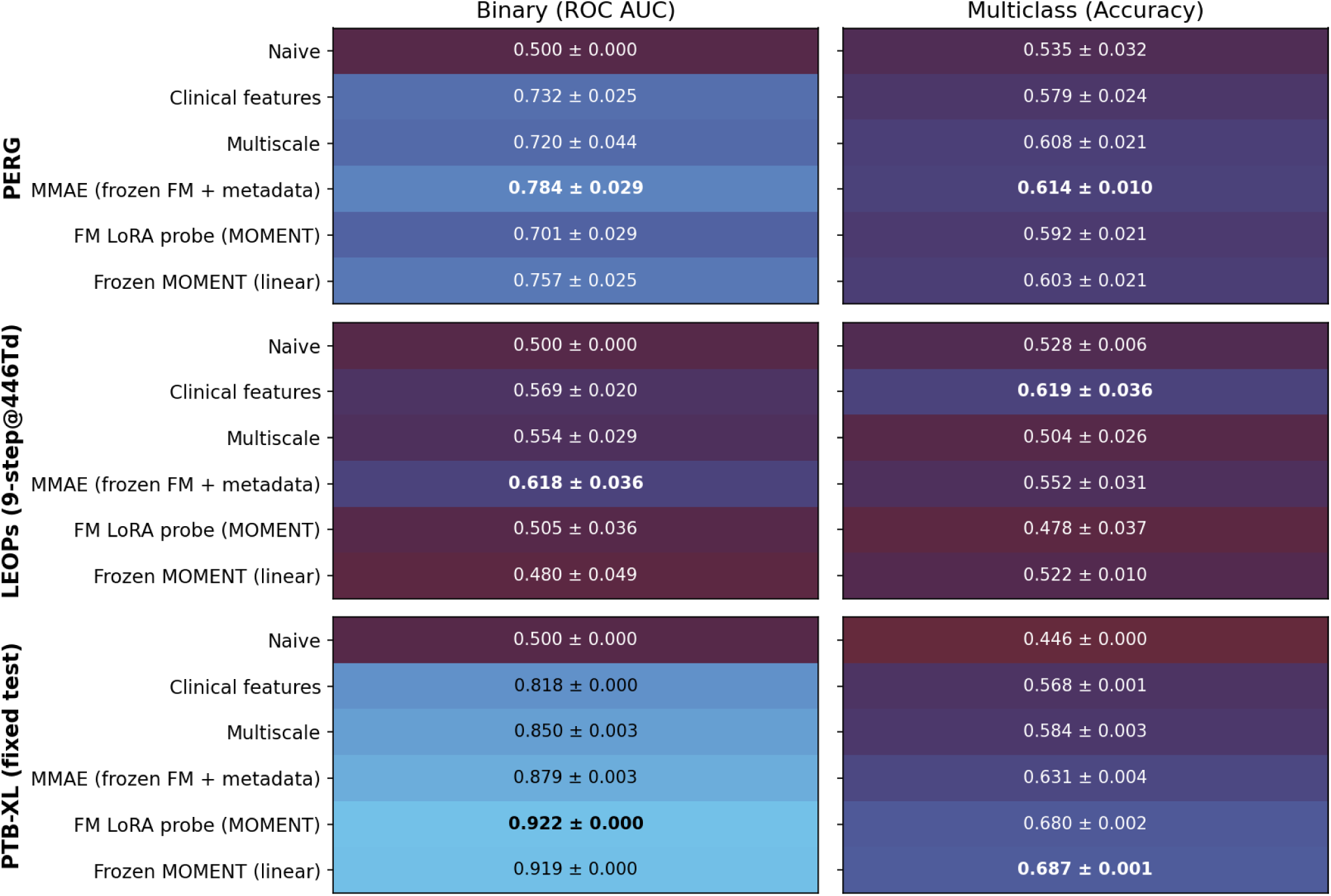
Classification performance for all finalist models on each dataset, against naive train set mean predictor. Values shown indicate Binary ROC AUC (left column) and multiclass accuracy (right column), represented as mean ± SEM across nested CV OOF samples.

### Classification Performance

#### PERG-IOBA

For PERG-IOBA’s binary classification task, the omnibus Friedman’s test was significant (X^2^ = 13.343, df = 5, p = 0.0204), and all tested models performed significantly better than the naive classifier (Figure 2, 3A). In contrast, no model was significantly better than the ISCEV clinical features put into a linear model (AUC=0.732 ± 0.025). The best single value was still a MMAE with frozen MOMENT embeddings (AUC=0.784 ± 0.029, Figure 3C), implying the multimodal model has some information the derived clinical components miss. Multiclass accuracy did not have a significant omnibus test (X^2^ = 3.988, df = 5, p = 0.5511). The best single value was still the MMAE with frozen MOMENT embeddings (accuracy = 0.614 ± 0.010).

**Figure 3:**
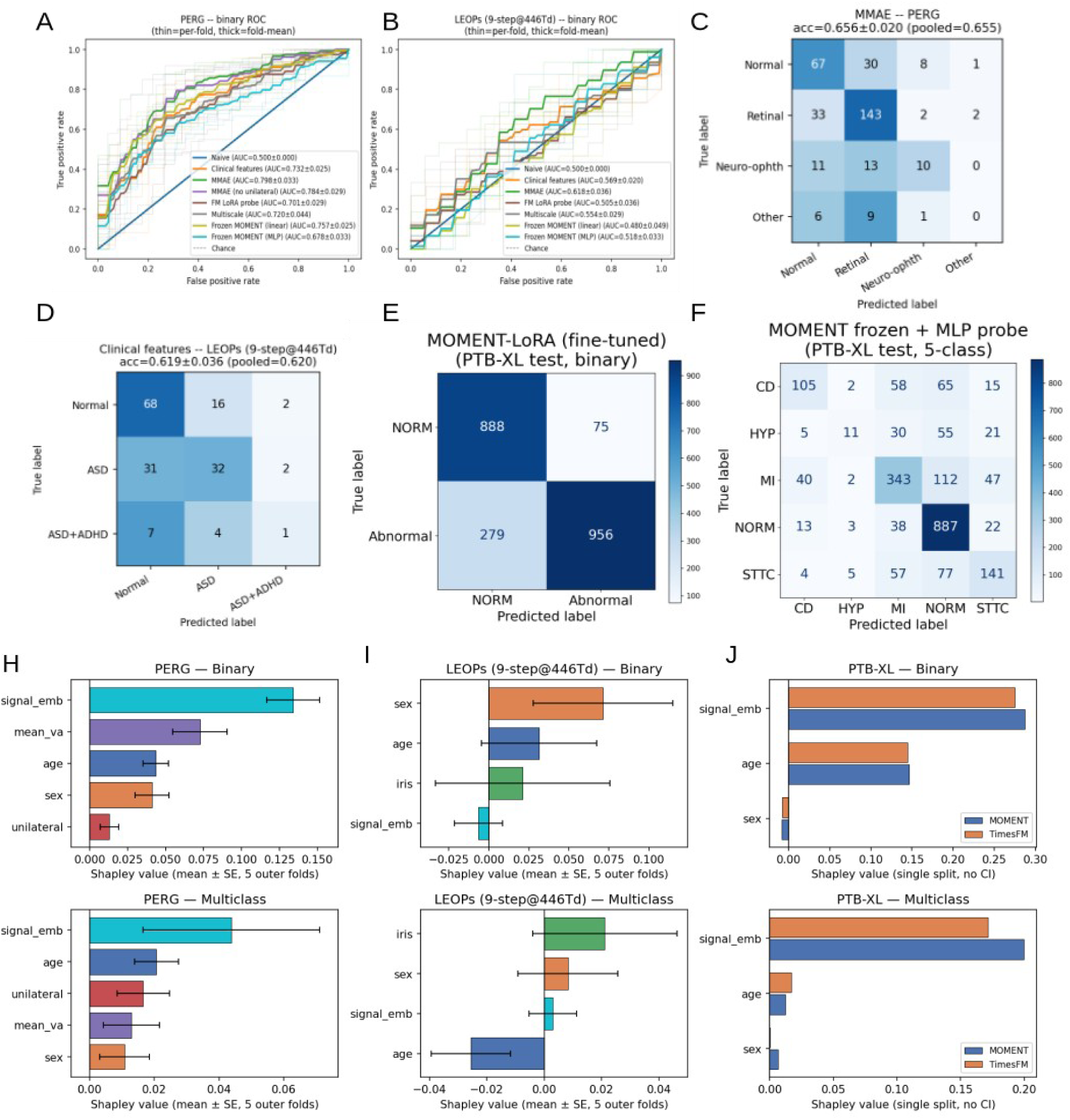
Classification Performance for Binary and Multiclass across models. **A and B** AUC-ROC curves for binary prediction performance from each finalist model, OOF predictions from nested 5-fold CV. MMAE performs significantly better than ISCEV PERG components even after masking laterality annotation. ASD was weakly predictable even with MMAE models, albeit better than ISCEV components. **C and D** Confusion matrices for the best multiclass models for PERG (**C**), and LEOPs (**D**) samples. **E** binary PTB-XL confusion matrix for LoRA fine-tuned MOMENT. **F** multiclass PTB-XL confusion matrix for frozen MOMENT with MLP probe. **H, I, J** Shapley analysis for each models’ MMAE variables.

When assessing the attribution of each modality to the MMAE performance, we found that the signal embedding was the strongest variable for binary and multiclass prediction (Figure 3H), although only significantly so in the binary. The other covariates all had positive attributions, and they seem to be genuinely synergistic given the raw performance delta between the MMAE with signal only (AUC = 0.725 ± 0.033) vs the full model (AUC = 0.788 ± 0.030). Despite this, all but unilateral were significantly better than chance for contribution scores to the binary task, which is likely due to unilateral being such a rare label in both train and test sets. Meanwhile, none of the attributions were significant in the multiclass task due to high variability.

Window-based Shapley attribution^35^ over the PERG trace (frozen MOMENT linear probe) showed attribution concentrated around the ISCEV N35/P50/N95 landmarks as well as in surrounding regions of the trace, especially near the tail of the recording (Figure 4).

**Figure 4:**
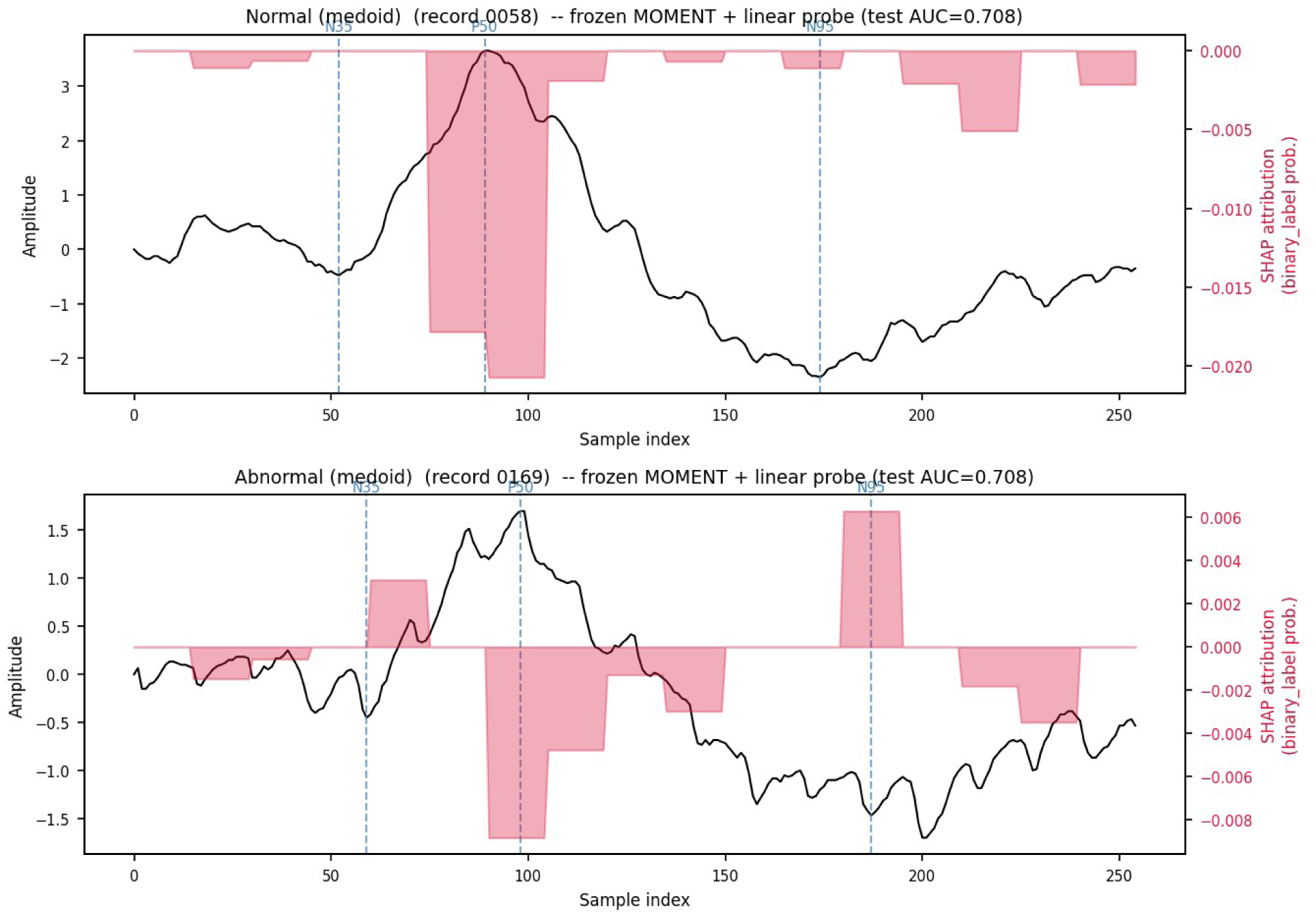
Window Shapley over normal and abnormal PERG traces.

#### LEOPs

Both the binary task of predicting ASD from typical development (X^2^ = 10.943, df = 5, p = 0.0525) and the multiclass adding ADHD annotations (X^2^ = 7.439, df = 5, p = 0.1900) were not significant at the level of the omnibus test (Figure 2, 3B). The best binary predictor was the frozen MMAE again (AUC=0.618 ± 0.036, Figure 3D), while the best multiclass model was actually the derived clinical features included with the dataset (AUC=0.619 ± 0.036). The failure of these results are interesting in light of the shapley analysis (Figure 3I). The binary task performance was heavily dependent on sex, which has a large distributional difference between TD and ASD (Table 1). The signal embeddings were barely better than chance. This failure seems to be a feature of the dataset, however, since the performance on PERG and PTB-XL were both significantly better than naive. These LEOPs results substantially underperform previously published ASD-classification approaches on related data (see Discussion).

#### PTB-XL

On PTB-XL, there were significant differences in both binary (X^2^ = 24.314, df = 5, p = 0.0002) and multiclass (X^2^ = 24.543, df = 5, p = 0.0002) omnibus tests. Unlike the other comparisons, the small error terms on the large evaluation set caused every post-hoc t-test to be significant. The strongest binary model was the MOMENT probe after LoRA fine-tuning (AUC=0.922 ± 0.000, Figure 3E), while the frozen linear probe was slightly better on multiclass accuracy (accuracy = 0.687 ± 0.001). In PTB-XL, the signal embedding was by far the variable with the highest Shapley attribution (Figure 3J), while age was helpful in binary and sex near chance in its utility.

### Age and Visual Acuity Prediction

**Figure 5:**
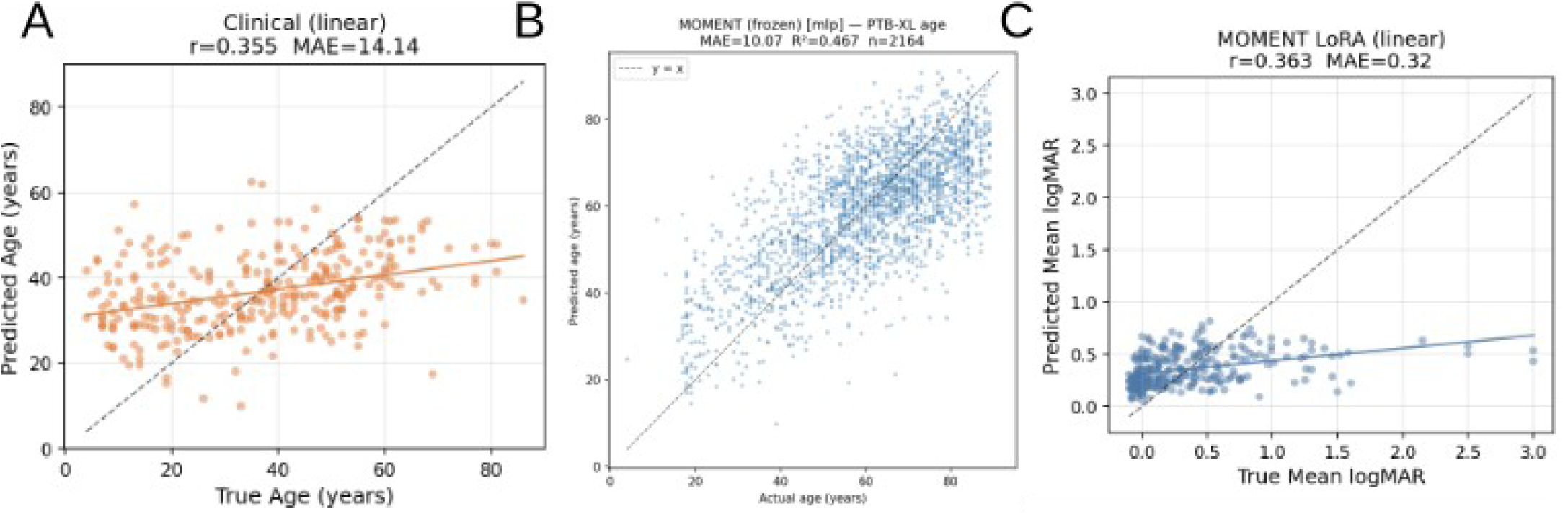
Performance on regression tasks. **A** Best PERG age prediction model, a linear model trained on the ISCEV components. MAE 14.14 years. **B** Best PTB-XL Age model, a linear probe from frozen MOMENT embeddings. **C** Best visual acuity model from PERG data, Fine-tuned MOMENT embeddings with a linear head. Significance not evaluated due to weak performance.

Age regression was weak with PERG data (Figure 5A). The best model was plain clinical features (MAE 14.14 ± 0.44 years), not significantly different from any FM-embedding model (frozen MOMENT MAE 14.34 ± 0.45, p=0.508 vs. clinical features), and only marginally better than a naive mean predictor (MAE 15.69 ± 0.25). FM embeddings were useful for age regression on PTB-XL (Figure 5B), however, where frozen MOMENT with an MLP head showed positive results (MAE 10.07 years, RZ=0.467, vs. naive MAE 14.07 years, ∼28% relative improvement). Visual acuity regression on PERG was also weak (best model: MOMENT LoRA with tied results for linear/MLP heads, Pearson *r* = 0.363, R^2^ ∼ 0.13, MAE 0.32 vs naive 0.35) and was never taken through nested cross-validation for formal testing due to the weak performance regardless of architecture (Figure 5C).

### Sample-Efficiency Scaling on PTB-XL

Subsampling PTB-XL’s training set (fixed 2,198-record test split; stratified resampling; binary AUC via linear probe on frozen embeddings) showed that most of the achievable performance is captured with as few as 300 training records (Figure 6). Frozen MOMENT reached AUC 0.877 ± 0.002 at N=300 versus 0.912 at the full N=17,418 (single run, no CI) — roughly 96% of full-dataset performance from 2.3% of the training data. Frozen TimesFM showed the same pattern (0.833 ± 0.004 at N=300 versus 0.887 at full N). MOMENT’s advantage over TimesFM held at every training-set size tested, but narrowed with more data (ΔAUC = 0.044 at N=300 vs. 0.025 at full N).

**Figure 6:**
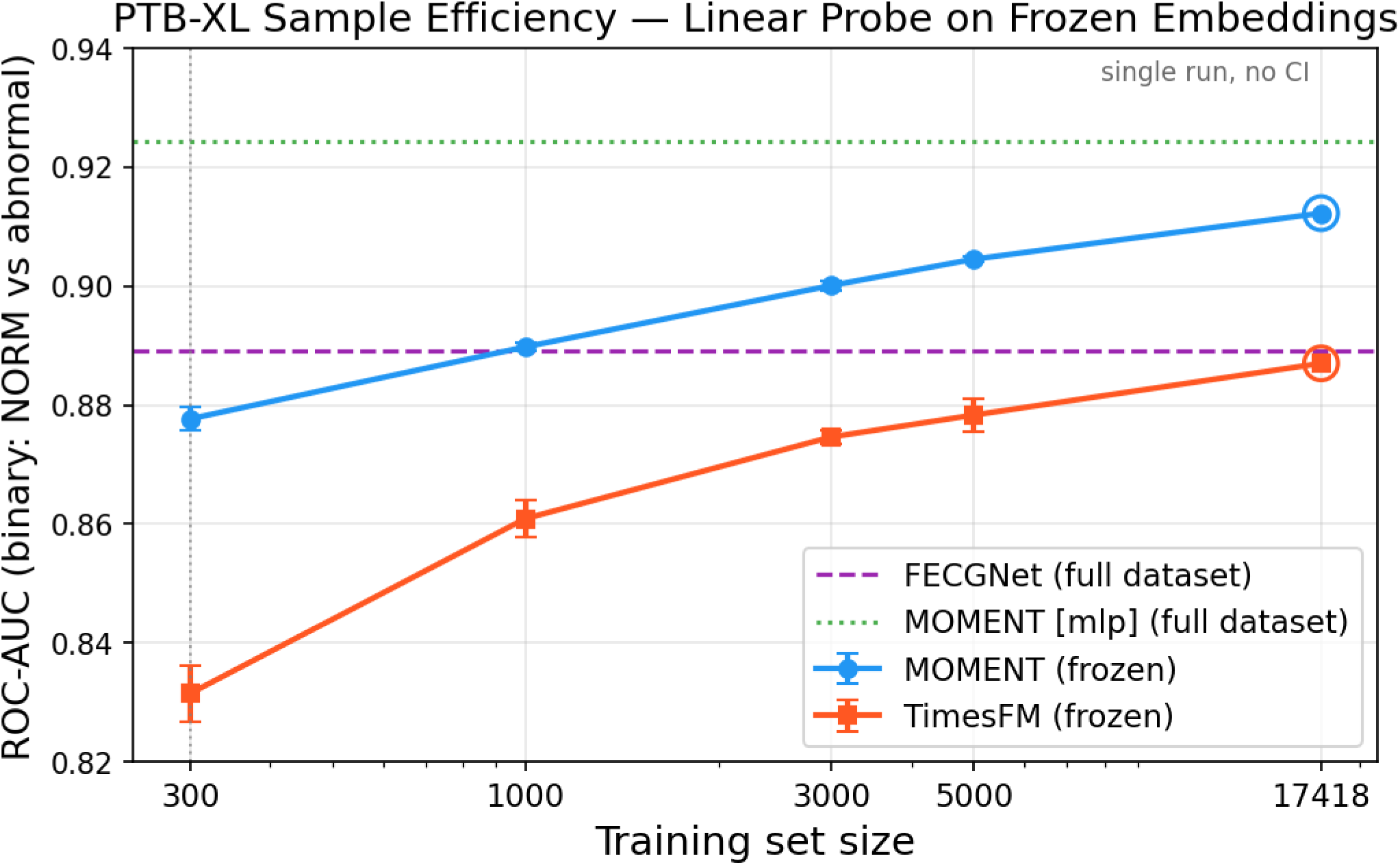
PTB-XL sample-efficiency scaling curve. Binary AUC vs. training-set size, frozen MOMENT/TimesFM linear probes, with FoundationalECGNet and MOMENT [mlp] full-dataset reference lines.

## IV. Discussion

### Strength of Time-Series Foundation Models and Current Clinical Components

Frozen FM embeddings were among the strongest approaches on most tasks. It may seem intuitive that models trained only within domain would trade generalization for internal validity and therefore perform better on a small dataset. Instead, the ability of these models to learn generic representations of time-series data transferred well into ERG in both datasets, and into ECG as well. Extracted clinical components were strong contenders across all tasks, implying that clinical practice converged on the most informative features of the trace even without modern methods. Our window Shapley analysis shows that deep learning models assign importance to those same components while also drawing signal from elsewhere in the trace directly supporting our second hypothesis.

This matters for several reasons. The strength of transfer learning from general-purpose FMs provides a usable baseline until ERG-specific pre-training corpora exist. The performance gain over extracted components, and the attribution of that gain to regions outside the standard measurement windows, suggests the standardization that made ERG comparable across laboratories also discarded usable information from within the full traces. FM embeddings were most effective alongside metadata covariates in the MMAEs, supporting multimodal fusion as a route to further gains in model accuracy and to improve their use in research or clinical settings. A further advantage of the FM approach is that a single model replaces multiple stimulus-specific models, meaning fine-tuning can draw on heterogeneous ERG data and may extend to stimuli for which no dedicated dataset yet exists. The masked pre-training objective of these FMs also improves the models’ robustness to nuisance variation across instruments, operators, and sites, helping to mitigate the problems that plagued early ERG exploration.

### Sample Efficiency and Scaling

Our PTB-XL subsampling analysis indicates that FM-based performance is maintained down to approximately 300 records, within 3-6 points of AUC across a 58-fold increase in training data, a size comparable to PERG-IOBA. This is the result with the broadest implications outside ERG, suggesting that transfer from large pre-trained time-series models is effective even at the small sample sizes currently available in ERG datasets. If it holds, ERG models approaching current ECG performance may be achievable at a fraction of the sample sizes the ECG literature required, without waiting for ERG datasets to reach comparable scale. MOMENT’s advantage over TimesFM persisted at every subsample size tested, from N=300 (Δ 0.044 AUC) to the full dataset (Δ 0.025 AUC), converging slightly as sample size grew.

### Architectural Comparisons

We evaluated architectures as published (e.g., MOMENT pre-trained externally and used frozen). Ideally, all models would be retrained in-house on the aggregated pre-training corpora used by MOMENT and TimesFM, allowing control over parameter count, distance of the pre-training modality from ERG (page views < traffic data < medical electrophysiology < ERG), and training decisions. This is costly to undertake without evidence that it would repay the compute, but our results support pursuing it. As we collect additional data, particularly alongside other measured modalities such as OCT rather than flat metadata values, this comparison will become more important. Models with far fewer parameters than MOMENT or TimesFM may be preferable before data reaches scale. Our PTB-XL results confirm that all tested architectures were correctly configured and capable of learning the task, and together they support frozen FMs as the strongest current strategy for ERG analysis using deep learning.

### Reproducibility of Published ASD Classification Results

Published reports describe strong ASD classification from bright-flash ERG, with AUCs of approximately 0.9 for VFCDM-based approaches ^24^ and 0.98 for UMAP-derived features ^25^. Our best models on this task reached AUC of 0.618 ± 0.036 (mean ± SE, 5 outer folds; no absolute confidence interval has been computed — the only interval available is a patient-level bootstrap on the delta vs. clinical features, +0.015 [95% CI −0.105, +0.131], not significant), and a model using demographic covariates alone (age and sex only, logistic regression, identical folds) reached AUC 0.589 ± 0.047, indicating that most of our performance derived from participant characteristics rather than the traces.

We investigated several mechanisms that could account for the discrepancy. Fitting the UMAP embedding on the full dataset before splitting rather than within each fold inflated performance relative to leakage-free evaluation but did not approach the published value (leakage-free/nested AUC 0.472 vs. leaky AUC 0.458, both near chance). Performing feature selection across all samples before cross-validation, as described for the VFCDM pipeline, similarly failed to reproduce the reported performance (feature-selection-order AUC: 9-step 446 Td none 0.621 / nested 0.586 / leaky 0.661; LA3 none 0.446 / nested 0.519 / leaky 0.506 — none approach the published ∼0.9). Splitting at the record rather than participant level, which would allow eyes or visits from the same individual to fall on both sides of a split, also did not close the gap (record-level leaky AUC up to 0.692 [random forest] vs. 0.597 under participant-grouped splitting — still well below 0.9). We additionally ran the published UMAP code as released on the data subset used in the original analysis (nested/as-written AUC 0.492; leaky pipeline variants attempted in their own execution order ranged 0.51–0.69; their notebook’s own cached output for the headline ∼0.98 could not be reconstructed from the released artifact, even when trying to copy their execution order). We therefore report that these results do not reproduce under patient-grouped, leakage-free evaluation, and that the mechanisms we tested do not account for the difference. Neither the VFCDM implementation nor its data preparation pipeline is publicly available, which limits how far reimplementation from the published description can settle the question.

These findings do not establish that the published results are incorrect, and remaining differences in cohort composition, label definition, or preprocessing between the released data versions could contribute. We note that the dataset from Anwar uses the 106 participant Flinders dataset, with 60/46 control/ASD samples. They do establish that the reported performance is not currently reproducible from public materials. Given that FM embeddings performed strongly on both PERG and ECG under identical evaluation, we consider it unlikely that our pipeline is uniquely deficient on this dataset.

The experience shaped our evaluation methodology. Early in this work we treated individual traces as the unit of analysis, on the assumption that a small amount of within-participant leakage would not materially change classification estimates. Testing that assumption showed otherwise, and we subsequently adopted pre-specified splits stored in the codebase alongside assertions and test suites that check for patient-level leakage at every stage of training and evaluation. All code and split definitions will be released upon acceptance.

### Limitations

Several limitations bound these conclusions. Both ERG datasets are small by machine learning standards, and PERG-IOBA in particular contains many diagnoses with few examples, requiring the coarser family groupings used here. We evaluated in two disease contexts and one control modality; whether these results extend to other ERG protocols or populations is untested. Architectures were compared as published rather than matched for parameter count or pre-training corpus, so differences between them confound model capacity with pre-training data. Performance on LEOPs was substantially attributable to demographic covariates, and the dataset’s composition may limit what any method can extract from the traces. We had no external validation cohort for either ERG dataset, and results are therefore cross-validated rather than externally validated. Finally, our reimplementations of FECGNet and VFCDM were derived from published descriptions without access to source code, and may differ from the original implementations in ways we cannot verify.

### Future Directions

As noted earlier, AI has already made a substantial mark on ophthalmology through color fundus photography and OCT models such as RETFound. ERG should provide a functional anchor that complements the structural information those models capture. We are now working toward methods for integrating FM embeddings across modalities, particularly in the data-scarce regimes that will persist until larger datasets become available. These models may be useful as a screening aid for ocular function, especially as devices such as RETeval make administering the tests easier for non-experts.

## Conclusions

Taken together, our results support time-series foundation models a strong approach for ERG inference. If similar scaling holds in ERG, models comparable to current ECG performance may be achievable with far fewer samples by transferring from public foundation models. We hope these findings encourage wider use of the electrophysiology data that already exists, and that others will share data to help push the frontier of what we can learn by looking through the eye.

## Author Contributions

**H.L.P.** — Conceptualization, Methodology, Software, Formal analysis, Investigation, Data curation, Visualization, Writing – original draft, Writing – review & editing

**C.B.G.** — Methodology, Writing – review & editing

**S.K.** — Formal analysis, Writing – review & editing

**J.D.W.** — Supervision, Funding acquisition, Resources, Writing – review & editing

## Data Availability

All data used in this project are publicly available. PERG-IOBA (https://physionet.org/content/perg-ioba-dataset/1.0.0/) and PTB-XL (https://physionet.org/content/ptb-xl/1.0.3/) were obtained from PhysioNet.org ^38^. LEOPs (https://data.mendeley.com/datasets/w3yx7hdds7/1) data were obtained from Mendeley Data.

## Code Availability

Code for all analyses, including pre-specified cross-validation splits, will be released publicly upon publication, subject to institutional licensing approval. Trained model weights will be made available on HuggingFace after publication.

## Ethics Statement

All data analyzed are de-identified, public datasets, and do not qualify as human subjects research. No new data were generated as part of this study.

